# A National Active Case-Finding Program for Tuberculosis in Prisons, Peru, 2024

**DOI:** 10.1101/2024.11.08.24317002

**Authors:** Esther Jung, Valentina A. Alarcón, Wilfredo Santos Solís Tupes, Tatiana Avalos-Cruz, Marco Tovar, Erika Abregu, Max Z. Yang, Jason R. Andrews, Moises A. Huaman

## Abstract

From January to September 2024, a national active case-finding program in Peru’s prisons screened more than 38,000 incarcerated persons for tuberculosis using chest radiography with automated interpretation and rapid molecular diagnostics, yielding a tuberculosis prevalence of 2,800 per 100,000 persons, with 11.4% of cases rifampicin-resistant; 42.5% of cases were subclinical.

**Article Summary Line:** A recent, nationwide mass screening program in Peru’s prisons using digital radiography with automated interpretation and molecular testing identified high tuberculosis prevalence and rates of rifampicin resistance, highlighting the need for active case-finding as an efficient strategy to identify undiagnosed TB burden in high-risk populations.

## The Study

Global tuberculosis (TB) incidence has slowly declined over the past decade, but prisons, especially in Latin America, have seen alarming increases in TB incidence over the same period (*1*). Overcrowding, poor ventilation, and diagnostic delays amplify tuberculosis transmission, such that incidence rates in this region are 27 times higher among persons deprived of liberty (PDL) compared to the general population (*2*). The rise in tuberculosis in Latin America’s prisons has more than offset reductions in TB in the general population, undermining progress towards the *End TB* goals (*3*).

To address this disproportionate burden, the World Health Organization (WHO) now recommends active case-finding (ACF) for TB in prisons (*4*). However, few standardized efforts have been made to implement nationwide screening for TB in prisons in low- and middle-income countries (LMICs). There remains a critical need for evidence-based, efficient implementation strategies that are sustainable in real-world settings. The Peruvian National TB Program (DPCTB) recently initiated a country-wide screening strategy using chest radiography (CXR) with computer-detection software, clinical evaluation and molecular rapid diagnostics in high TB-burdened prisons. We sought to evaluate the programmatic yield of this initiative and assess how each screening component contributed to identifying tuberculosis cases.

Peru has a population of 34 million inhabitants and 96,805 prisoners (*5,6*). In 2022, national TB incidence was estimated to be 151 cases per 100,000 person-years (*6*), while incidence among PDL has been estimated to be 4,194 cases per 100,000 person-years (*7*). In 2023, DPCTB implemented an active case finding program in 12 male, 1 co-ed, and 5 female prisons chosen for size, TB burden, and convenience (Figure 1).

**Figure 1.**
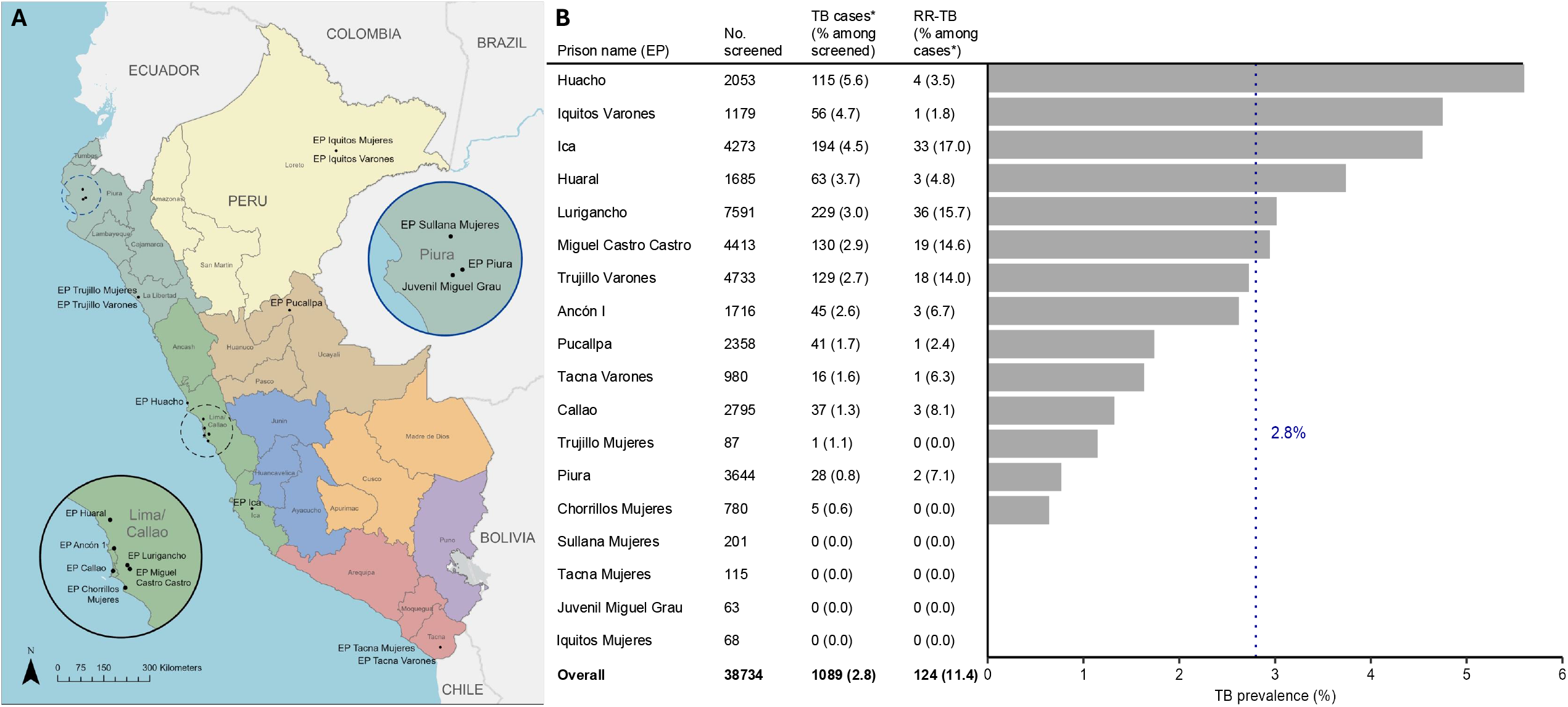
Panel A - Map of 18 prisons included in screening procedures, insets magnify Lima/Callao and Piura districts. Panel B - Bar diagram displaying tuberculosis prevalence and rifampicin-resistance (RR-TB) by prison, sorted by descending TB prevalence. Dotted line shows overall TB prevalence across all prisons (2.8%). *TB case defined as any positive or trace Xpert result. EP, Establecimiento Penitenciario (Penitentiary Establishment); Juvenil Miguel Grau, Centro Juvenil de Diagnóstico y Rehabilitación Miguel Grau; Mujeres, women’s prison, RR-TB, rifampicin-resistant TB; TB, tuberculosis, Varones, men’s prison.

Programmatic data from PDL ≥ 18 years old who were not being treated for active TB and received a CXR screening between January and September 2024 were included in this analysis. The screening team consisted of a physician, nurse, and radiology technician; three largest prisons included additional lab personnel. Each participant was interviewed to collect demographic, clinical, and TB-symptom information. All participants were screened with a portable digital CXR machine, and CXR were evaluated with Computer-Aided Detection for Tuberculosis (CAD4TB) software version 7.0. CAD4TB assigns a score between 1 and 100 based on radiological abnormalities suggestive of a TB diagnosis. All participants then underwent clinical evaluation by a physician.

Participants with a CAD4TB score ≥□40 (considered “abnormal”) were asked to produce a sputum sample. Participants with a CAD4TB score < 40 (considered “normal”) were only asked to produce a sputum sample if the physician suspected TB based on symptoms and/or evaluation. A molecular rapid diagnostic test, Xpert MTB/RIF Ultra assay (Cepheid, Sunnyvale; hereinafter Xpert), was performed on all but 308 sputum samples, which were excluded from analysis. We defined a TB case as any positive or trace sputum Xpert result. All individuals with confirmed TB were provided free treatment.

Between January and September of 2024, DPCTB screened 38,734 participants meeting inclusion criteria: 7,291 (18.8%) had sputum collected, and 6,873 (94.3%) of samples produced valid Xpert results (Figure 2). To evaluate the association between demographic and clinical characteristics and TB, we used multivariable logistic regression with fixed effects for prison to estimate crude (OR) and adjusted (AOR) odds ratios with their respective 95% confidence intervals. We calculated the sputum positivity and proportion of TB cases detected by combinations of symptom screen and CAD4TB results (Table 2). R (version 4.4.1) was used for statistical analyses.

**Table 1.** Multivariable logistic regression analysis of demographic characteristics and risk factors for tuberculosis among screened PDL.

| Risk factor | n | TB Confirmed (%) | OR (95% CI) | *P* value | AOR (95% CI) | *P* value |
| --- | --- | --- | --- | --- | --- | --- |
| Age |  |  |  |  |  |  |
| 18-29 | 11193 | 382 (3.4) |  |  |  |  |
| 30-44 | 17557 | 486 (2.8) | 0.75 (0.65, 0.87) | <.001 | 0.68 (0.59, 0.78) | <.001 |
| 45-60 | 7708 | 169 (2.2) | 0.58 (0.48, 0.70) | <.001 | 0.54 (0.45, 0.65) | <.001 |
| ≥ 60 | 2274 | 52 (2.3) | 0.61 (0.45, 0.81) | <.001 | 0.58 (0.42, 0.77) | <.001 |
| Sex |  |  |  |  |  |  |
| Male | 37227 | 1081 (2.9) | 5.01 (1.60, 30.3) | .024 | 3.78 (1.20, 22.94) | .062 |
| Female | 1507 | 8 (0.5) |  |  |  |  |
| TB History |  |  |  |  |  |  |
| Yes | 6232 | 397 (6.4) | 2.85 (2.50, 3.24) | <.001 | 2.75 (2.41, 3.13) | <.001 |
| No | 32502 | 692 (2.1) |  |  |  |  |
| TB Contact | 15459 | 556 (3.6) | 2.15 (1.77, 2.59) | <.001 | 1.90 (1.56, 2.30) | <.001 |
| Peru Origin | 36596 | 1054 (2.9) | 1.80 (1.30, 2.58) | .001 | 1.67 (1.20, 2.41) | .004 |

Data are presented as no. (%) unless otherwise indicated. CI, confidence interval; PDL, persons deprived of liberty;
TB, tuberculosis; OR, odds ratio; AOR, adjusted odds ratio; PDL, persons deprived of liberty.

**Table 2.**
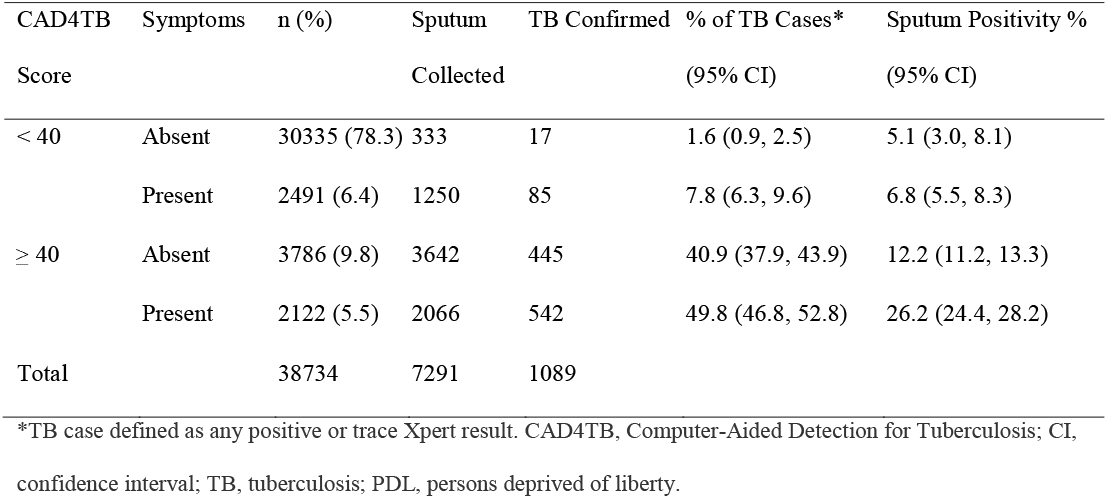
Predictive value of Computer-Aided Detection for Tuberculosis Score, symptom screen, and sputum collection among screened PDL.

**Figure 2.**
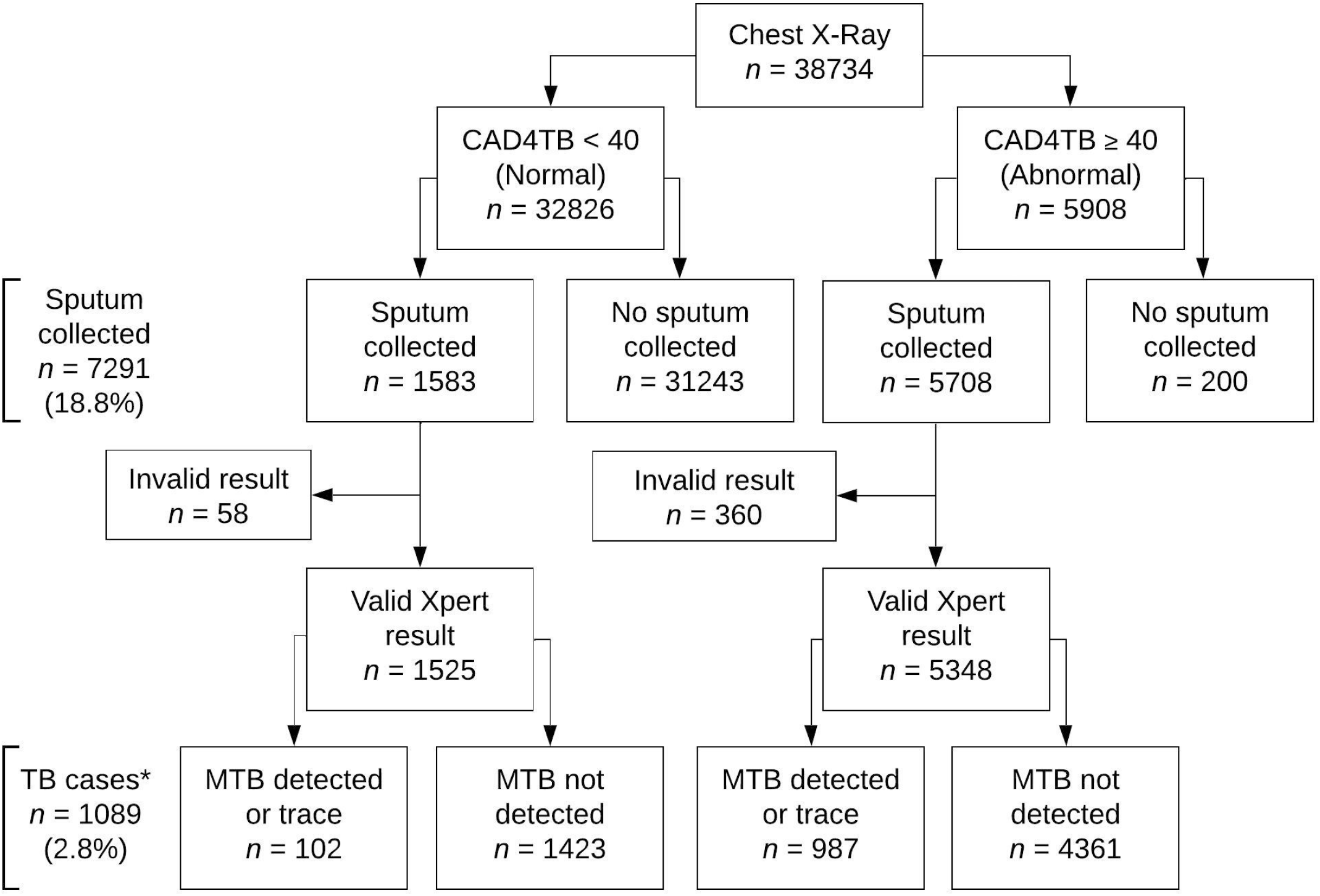
Flowchart for tuberculosis screening algorithm across 18 study prisons among included participants. *TB case defined as any positive or trace Xpert result. CAD4TB, Computer-Aided Detection for Tuberculosis; MTB, Mycobacterium tuberculosis; PDL, persons deprived of liberty; TB, tuberculosis.

Most study participants were male (96%), born in Peru (94%), and median age was 35 years old (IQR 27-43). Previous TB history was reported among 16% of participants, and 40% reported prior contact with TB (Table 1).

Among all participants, 1,089 (2.8%) were diagnosed with TB. Prevalence ranged from 0% in several small prisons (<250 PDL) to 5.6% among 2,053 PDL in Huacho. All five female prisons had a prevalence under 1.5%, while 8 of the 12 male prisons had TB prevalence over 2%. 11.4% (124/1089) of Xpert-positive samples tested positive for rifampicin resistance (RR-TB), with four prisons recording RR-TB prevalence among cases of over 10% (Figure 1).

5,908 (15.3%) participants had abnormal CAD4TB scores, and 4,613 (11.9%) reported symptoms in the preceding two weeks. Xpert positivity varied substantially according to symptoms and x-ray abnormality. Among individuals with symptoms but normal scores, 6.8% had positive Xpert, representing 7.8% of TB cases detected. Among individuals with abnormal scores but no symptoms, 12.2% had positive Xpert, representing 40.9% of TB cases. Xpert was positive in 26.2% of those with symptoms and abnormal scores, who comprised 49.8% of TB cases (Table 2).

Odds of TB were higher among participants with a reported history of TB (AOR: 2.79, 95% CI, 2.44-3.19) and contact with TB (AOR: 1.90, 95% CI: 1.55-2.31) (Table 1). Odds of RR-TB were higher among cases with a reported history of TB (AOR: 1.95, 95% CI, 1.32-2.91) (Supplementary Table 1).

## Conclusion

There is a critical need for successful, scalable models for ACF in LMIC prisons, where estimates suggest that nearly half of cases go undetected annually (*7*). In Peru, a mass screening strategy led by mobile health teams using CXR with automated interpretation screened over 38,000 PDL in eight months, more than one-third of the national incarcerated population, making it one of the largest scale implementations of ACF based in molecular testing in LMIC prisons reported to date. The prevalence of undiagnosed TB was 2,800 per 100,000, with 11.4% of RR-TB. More than 40% of individuals with Xpert-confirmed TB lacked symptoms. These findings illustrate the importance of systematic screening in prisons, including among individuals without TB symptoms.

This study also presents the first nationally representative prevalence estimates of TB burden in Peru’s carceral system based in molecular testing. Since 2020, Peru has remained on the WHO’s list of the 30 countries with the highest levels of drug-resistant TB, and it is estimated that 8.3% of new TB diagnoses every year are drug-resistant (*6,8*). Our study detected a high prevalence of TB (2,800 per 100,000), and a very high proportion (11.4%) of RR-TB. The prevalence estimate was almost certainly an underestimate for these prisons, as sputum was only collected from participants with x-ray abnormalities, symptoms, or clinical suspicion of TB. Including clinical inference as a criterion would likely capture additional TB. However, this study was limited to 18 prisons selected for high TB-burden and convenience, hindering generalizability to other prisons in Peru.

A hidden threat in high-transmission settings like prisons in LMICs is subclinical TB. Previous estimates have shown that asymptomatic TB accounts for a considerable proportion of active disease and overall transmission in both the general and incarcerated population (*9-12*). We found 42.5% of all TB cases in this study to be subclinical, likely an underestimate due to the inclusion of symptoms in the screening algorithm criteria, suggesting that symptom-only case-finding strategies would delay detection or miss a significant fraction of active TB among PDL.

Our data demonstrate that active case-finding can identify a large reservoir of undiagnosed tuberculosis and can be performed efficiently at scale in prisons using mobile radiography with automated interpretation led by teams of health professionals. Such strategies require incorporation into national TB programs with sustainable financing to roll out and maintain their implementation at scale in LMIC prisons, where the burden of TB demands urgent attention.

## Data Availability

All data produced in the present study are available upon reasonable request to the Peruvian National TB Program.

## Acknowledgements

We thank the Peruvian National TB Program team for providing data and assistance during this study.

## Biographical Sketch

Ms. Jung is a research program coordinator at Stanford University School of Medicine. Her research interests include environmental and infectious disease epidemiology in marginalized populations.

## References

1. Walter KS, Martinez L, Arakaki-Sanchez D, Sequera VG, Sanabria GE, Cohen T, Ko AI, García-Basteiro AL, Rueda ZV, López-Olarte RA, Espinal MA, Croda J, Andrews JR. The escalating tuberculosis crisis in central and South American prisons. The Lancet. 2021;397(10284),1591–1596. DOI

2. Cords O, Martinez L, Warren JL, O’Marr JM, Walter KS, Cohen T, Zheng J, Ko AI, Croda J, Andrews JR. Incidence and prevalence of tuberculosis in incarcerated populations: A systematic review and meta-analysis. The Lancet Public Health. 2021;6(5). DOI

3. World Health Organization. The End TB Strategy: Global strategy and targets for tuberculosis prevention, care and control after 2015. 2014 [cited 2024 Aug 23]. https://www.who.int/publications/i/item/WHO-HTM-TB-2015.19

4. World Health Organization. Consolidated guidelines on tuberculosis: Module 2: screening – systematic screening for tuberculosis disease. 2021 [cited 2024 Aug 2]. http://www.ncbi.nlm.nih.gov/books/NBK569338/

5. World Prison Brief. Peru. 2024 Apr 30 [cited 2024 Jul 13]. https://www.prisonstudies.org/country/peru

6. World Health Organization. TB profile. 2022 [cited 2024 Aug 13]. https://worldhealthorg.shinyapps.io/tb_profiles/?_inputs_&entity_type=%22country%22&iso2=%22PE%22&lan=%22EN%22

7. Martinez L, Warren JL, Harries AD, Croda J, Espinal MA, Olarte RAL, Avedillo P, Lienhardt C, Bhatia V, Liu Q, Chakaya J, Denholm JT, Lin, Y, Kawatsu L, Zhu L, Horsburgh CR, Cohen T, Andrews, JR. Global, regional, and national estimates of tuberculosis incidence and case detection among incarcerated individuals from 2000 to 2019: A systematic analysis. The Lancet Public Health. 2023;8(7), e511.–e519. DOI

8. World Health Organization. Global Tuberculosis Report 2023. 2023 Nov 7 [cited 2024 Jul 23]. https://www.who.int/teams/global-tuberculosis-programme/tb-reports/global-tuberculosis-report-2023

9. Frascella B, Richards AS, Sossen B, Emery JC, Odone A, Law I, Onozaki I, Esmail H, Houben, RMGJ. Subclinical Tuberculosis Disease — A Review and Analysis of Prevalence Surveys to Inform Definitions, Burden, Associations, and Screening Methodology. Clinical Infectious Diseases. 2020;73(3), e830.–e841. DOI

10. Kendall EA, Shrestha S, Dowdy DW. The Epidemiological Importance of Subclinical Tuberculosis. A Critical Reappraisal. Am J Respir Crit Care Med. 2021 Jan 15;203(2):168–174. DOI.

11. Puma, D., Yuen, C. M., Millones, A. K., Brooks, M. B., Jimenez, J., Calderon, R. I., Lecca, L., Becerra, M. C., & Keshavjee, S. (2023). Sensitivity of Various Case Detection Algorithms for Community-based Tuberculosis Screening. Clinical Infectious Diseases, 76(3), e987.–e989. DOI

12. Puma, D., Geadas, C., Calderon, R. I., Yuen, C. M., Jiménez, J., Córdova, M., Martínez, B., Peinado, J., Lecca, L., & Tovar, M. (2023). Active case-finding for TB among incarcerated women in Peru. The International Journal of Tuberculosis and Lung Disease, 27(10), 784. DOI

