## Supplemental Table for "A National Active Case-Finding Program for Tuberculosis in Prisons, Peru, 2024"

**Supplemental Table.** Multivariable logistic regression analysis of demographic characteristics and risk factors for rifampicin-resistant TB among PDL with confirmed TB.

| Risk factor | TB Confirmed (n) | RR-TB (%) | OR (95% CI) | *P* value | AOR (95% CI) | *P* value |
|---|---|---|---|---|---|---|
| Age | | | | | | |
| 18-29 | 382 | 40 (10.5) | | | | |
| 30-44 | 486 | 63 (13.0) | 1.31 (0.86, 2.04) | .215 | 1.18 (0.76, 1.85) | .462 |
| 45-60 | 169 | 16 (9.5) | 1.07 (0.56, 1.99) | .830 | 0.97 (0.50, 1.82) | .928 |
| ≥ 60 | 52 | 5 (9.6) | 1.44 (0.46, 3.72) | .482 | 1.32 (0.42, 3.45) | .597 |
| Sex | | | | | | |
| Male | 1081 | 123 (11.4) | 0.20 (0.01, 5.14) | .260 | 0.15 (0.01, 3.99) | .192 |
| Female | 8 | 1 (12.5) | | | | |
| TB History | | | | | | |
| Yes | 397 | 65 (16.4) | 2.00 (1.36, 2.96) | <.001 | 1.96 (1.32, 2.91) | .001 |
| No | 692 | 59 (8.5) | | | | |
| TB Contact | 556 | 56 (10.1) | 0.61 (0.32, 1.11) | .119 | 0.59 (0.31, 1.08) | .103 |
| Peru Origin | 1054 | 122 (11.6) | 2.72 (0.80, 17.02) | .176 | 2.25 (0.65, 14.21) | .279 |

Data are presented as no. (%) unless otherwise indicated. CI, confidence interval; PDL, persons deprived of liberty; TB, tuberculosis; RR-TB, rifampicin-resistant tuberculosis; OR, odds ratio; AOR, adjusted odds ratio; PDL, persons deprived of liberty.
